# Novel Therapeutic Strategy for Orthostatic Hypotension Using Deep Brain Stimulation

**DOI:** 10.64898/2026.04.14.26350914

**Authors:** Fumiyasu Yamasaki, Masato Seike, Takayoshi Hirota, Takayuki Sato

**Author notes:** Address for correspondence: Fumiyasu Yamasaki, MD Department of Clinical Laboratory, Kochi Medical School, Nankoku, 783-8505, Japan (ext 3431).

## Abstract

**Background:** Deep brain stimulation (DBS) is a treatment option for Parkinson disease (PD). However, the effect of DBS on the arterial pressure (AP) remains unexplored. We aimed to develop an artificial baroreflex system for treating orthostatic hypotension (OH) due to central baroreflex failure in patients with PD. To achieve this, we developed an appropriate algorithm after estimating the dynamic responses of the AP to DBS using a white noise system identification method.

**Methods:** We randomly performed DBS while measuring the AP tonometrically in 3 trials involving 3 patients with PD treated with DBS. We calculated the frequency response of the AP to the DBS using a fast Fourier transform algorithm. Finally, the feedback correction factors were determined via numerical simulation.

**Results:** The frequency responses of the systolic AP to random DBS were identifiable in all 3 trials, and the steady state gain was 8.24 mmHg/STM. Based on these results, the proportional correction factor was set to 0.12, and the integral correction factor was set to 0.018. The computer simulation revealed that the system could quickly and effectively attenuate a sudden AP drop induced by external disturbances such as head-up tilting.

**Conclusion:** An artificial baroreflex system with DBS may be a novel therapeutic approach for OH caused by central baroreflex failure.

## Main Text

In patients with Parkinson disease (PD), autonomic failure often manifests as severe orthostatic hypotension (OH) due to arterial baroreflex failure in the advanced stages of the disease (1–4). While some medications or non-pharmacological therapies may provide relief from symptoms, they are not satisfactory solutions for OH and may have adverse effects (5,6). However, some exploratory clinical studies on deep brain stimulation (DBS) (7–10), a non-pharmacological therapy, report that it may be effective in OH (11–14). These findings suggest that DBS often has a pressor effect through sympathoexcitation in the brain, indicating that the baroreflex failure that occurs in some patients with PD is of a central type. This suggestion is associated with the applicability of bionic technology as a novel therapeutic strategy for treating OH in PD.

In a previous study utilizing bionic technology, we developed an artificial baroreflex system with an electrical stimulator of sympathetic nerves via an epidural catheter placed at the level of the lower thoracic spinal cord in patients with high cervical spinal cord injury (15,16). The computer system of the artificial baroreflex system served as the artificial vasomotor center, automatically computing the frequency of a pulse train (STM) required to stimulate the sympathetic nerves in response to arterial pressure (AP) changes and execute the appropriate stimulation of the sympathetic nerves.

The objective of the present study was to develop a feedback control algorithm for an artificial vasomotor center to restore the arterial baroreflex function in patients with PD and severe OH due to central-type baroreflex failure. In this study, we delineated the dynamic response of the AP to DBS and devised a feedback control algorithm for the vasomotor center of an artificial baroreflex system.

## METHODS

### Theoretical Considerations

As previously described (15,16), the operating principle of an artificial vasomotor center to restore arterial baroreflex function in central-type baroreflex failure is based on a negative feedback mechanism (Figure 1). The instantaneous change in the AP is negatively fed back to an artificial vasomotor center, which automatically performs DBS to minimize the effect of an external perturbation, such as standing up from the supine position, on the AP (P_ex_). The operating algorithm for the STM that drives the DBS generator is represented by the transfer function H_1_(*f*) in the frequency domain. The H_1_(*f*) is designed according to the principles of classical control theory, i.e., feedback correction with proportional and integral gain factors. In this theoretical framework, the command directed to the DBS generator for correction or control is calculated as the sum of the proportional and integral terms. The proportional term is the product of the proportional gain factor K_p_ and the current instantaneous change in AP, whereas the integral term is the product of the integral gain factor K_i_ and the cumulative value of past instantaneous changes in AP over time. Consequently, as shown in Figure 1, the frequency-domain STM, which is the output of a controller, i.e., the artificial vasomotor center, is calculated as follows:

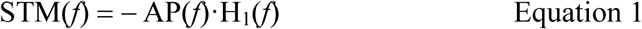

where H_1_(*f*) = K_p_ + K_i_ / 2πfj, the variable *f* represents the frequency, and *j* is the imaginary unit. On the other hand, AP in the frequency domain is expressed as follows:

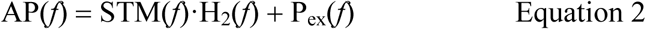

where H_2_(*f*) is the frequency response of the AP to the STM in the effector.

From Equations 1 and 2, the AP is estimated in the frequency domain as follows:

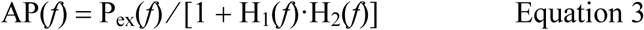

which means that the effect of an orthostatic challenge, P_ex_, on AP is attenuated to 1 / [1 + H_1_(*f*)·H_2_(*f*)] by the implementation of the artificial baroreflex system. To optimize the functional capacity of the artificial baroreflex system, H_2_(*f*) was first estimated from the observed measurements and then H_1_(*f*) was designed using a numerical simulation.

**Figure 1.**
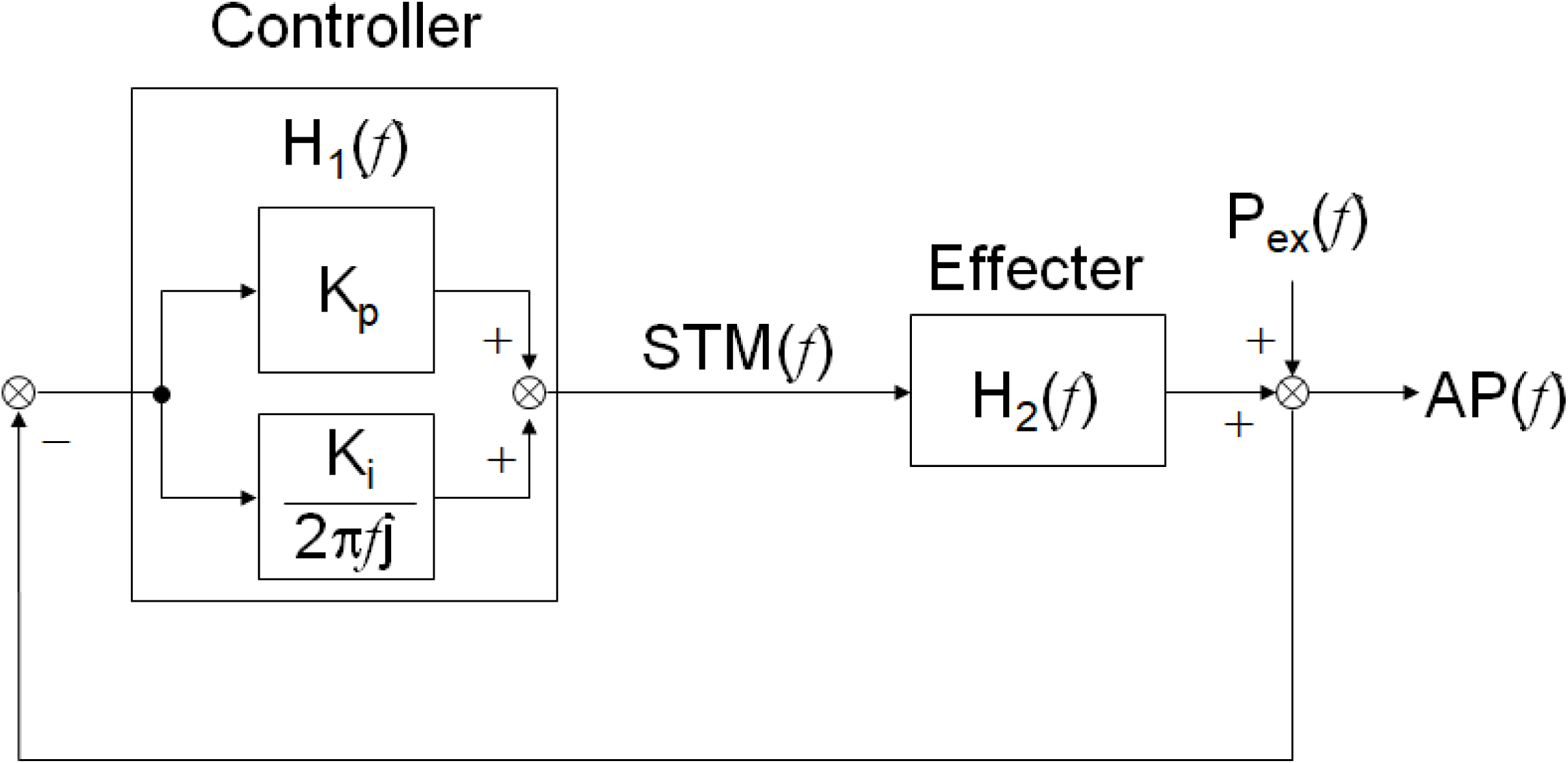
Block diagram of a servo model in an artificial vasomotor center. The artificial vasomotor center automatically computes the frequency (STM) of a pulse train to stimulate the deep brain, while simultaneously sensing the change in AP. H_1_(*f*) denotes a transfer function for the controller functioning as an artificial vasomotor center. H_2_(*f*) is a transfer function showing the dynamic response of AP to DBS. The overall transfer function of the artificial vasomotor center is given by H_1_(*f*)·H_2_(*f*). Therefore, the effect of an external disturbance (P_ex_) on AP is attenuated to 1 / [1 + H_1_(*f*)·H_2_(*f*)].

### Subjects

Fourteen patients with PD (mean age: 64 ± 11 years; seven male) who had undergone bilateral subthalamic nucleus DBS participated in this study. The mean disease duration was 14 ± 6 years. The mean Hoehn and Yahr stage at the time of the trial was 4.0 ± 1.0. Thirteen patients were taking levodopa, 9 were taking dopamine agonists, and seven were taking other medications, including amantadine. Five patients presented with hypertension, two had diabetes mellitus, and two had cardiovascular disease. None of the participants exhibited frequent ectopic beats or atrial fibrillation. The Leksell SurgiPlan navigation system (Elekta Instrument AB, Stockholm, Sweden) was used to implant the DBS electrodes in the subthalamic nuclei (STN). All patients underwent implantation of quadripolar Model 3387-28 electrodes, comprising four contacts at the extremities of the electrode. The electrodes were connected to Soletra, ActivaSC, or ActivaSC pulse generators (Medtronic Inc., Minneapolis, MN, USA). Before the commencement of the study, all participants were provided with relevant information and, where appropriate, were given the opportunity to consent to their participation. The study protocol was approved by the local ethics committee of Izumino Hospital (approval number 1-20120712) and adhered to the ethical standards of the Committee on Human Experimentation and the Declaration of Helsinki.

### Measurement and Responder for the Stimulation

Data acquisition was conducted in a quiet, temperature-controlled room at 15:00, at least two hours after the participants had eaten lunch. The room temperature was 23°C. AP was measured tonometrically using a JENTOW device (Colin, Komaki, Japan) with concurrent electrocardiographic (ECG) recordings. Following a 10-minute stabilization period, AP and ECG were recorded continuously in the supine position. Subsequently, a hemilateral test stimulus was applied for 20 seconds. The frequency and current of the stimuli were set to the highest possible levels via the most distal pole without causing any discomfort. A subject whose blood pressure increased by 10 mmHg or more was designated as a responder. Finally, a binary, white noise-like, random STM of “on” or “off” was performed unilaterally for 12 minutes in the responders with the same frequency and current as the test stimulus. (Figure 2).

**Figure 2.**
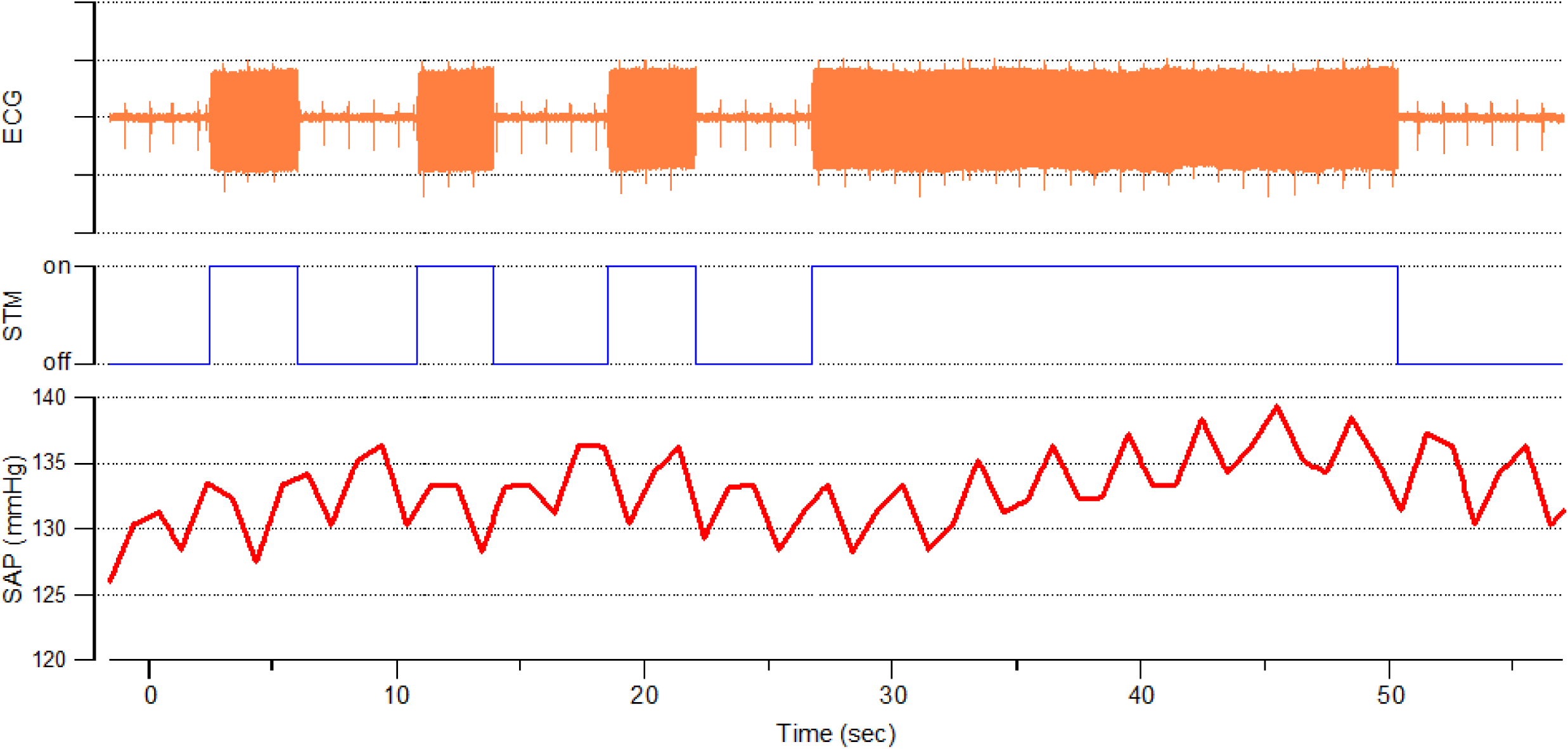
The data line of the ECG, STM, and SAP (systolic AP). The unilateral DBS generator stimulation was randomly turned on or off. We manually created an STM line using an electrical artefact on the ECG line. This STM line was then used to calculate the transfer function from DBS to systolic AP.

### Estimation of Transfer Function from STM to AP

The dynamic nature of the AP response to STM, i.e., H_2_(*f*), was estimated in the responders using a white noise method for system identification. As described previously (15–18), for each patient, H_2_(*f*) was estimated from the time-series data of STM and systolic AP based on a fast Fourier transform algorithm using Igor Pro version 3.16 (WaveMetrics, Lake Oswego, USA). Then, the average H_2_(*f*) in the responder trials was calculated. To parametrically characterize the averaged H_2_(*f*), it was approximated using an iterative nonlinear least-squares fit to a model for a first-order low-pass filter as follows:

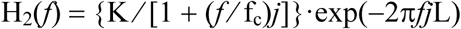

where K is the steady-state gain, fc is the cut-off frequency, and L is the pure delay. The final parametric estimate for H_2_(*f*) is used in a numerical simulation to design H_1_(*f*).

### Design of Artificial Vasomotor Center

The final parametric estimate for H_2_(*f*) was substituted into Equation 2, and the instantaneous AP response to P_ex_ was numerically simulated using time domain analysis, assuming a sudden stepwise decrease with an amplitude of 10 mmHg imposed by the external perturbation P_ex_ on the negative feedback system. As described previously, the effect of changing the feedback correction factors K_p_ and K_i_ by 1 on the AP response under closed-loop conditions of the artificial baroreflex system was evaluated using a convolution integral algorithm as follows:

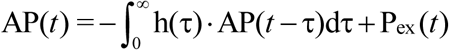

where h(τ) is the impulse response function of the total open-loop transfer function, H_1_(*f*)–H_2_(*f*). Finally, correction factors were identified that effectively and rapidly attenuated the effect of P_ex_ on AP.

## RESULTS

A total of 19 test stimuli on unilateral DBS were administered to 14 patients, comprising 12 tests on the right side and seven on the left side. The tests were performed 36 ± 36 months after the DBS surgery. The DBS settings used for the test stimuli are provided in Table 1. Of the original pulse generator trials, eight used monopolar stimulation and 11 used bipolar stimulation. All the settings were changed to monopolar stimulation. The amplitude changed from 2.6 ± 0.5 mA to 2.7 ± 0.7 mA, and the stimulation rate was from 136 ± 13 Hz to 143 ± 12 Hz. The pulse width is maintained at 90 ms.

**Table 1.**
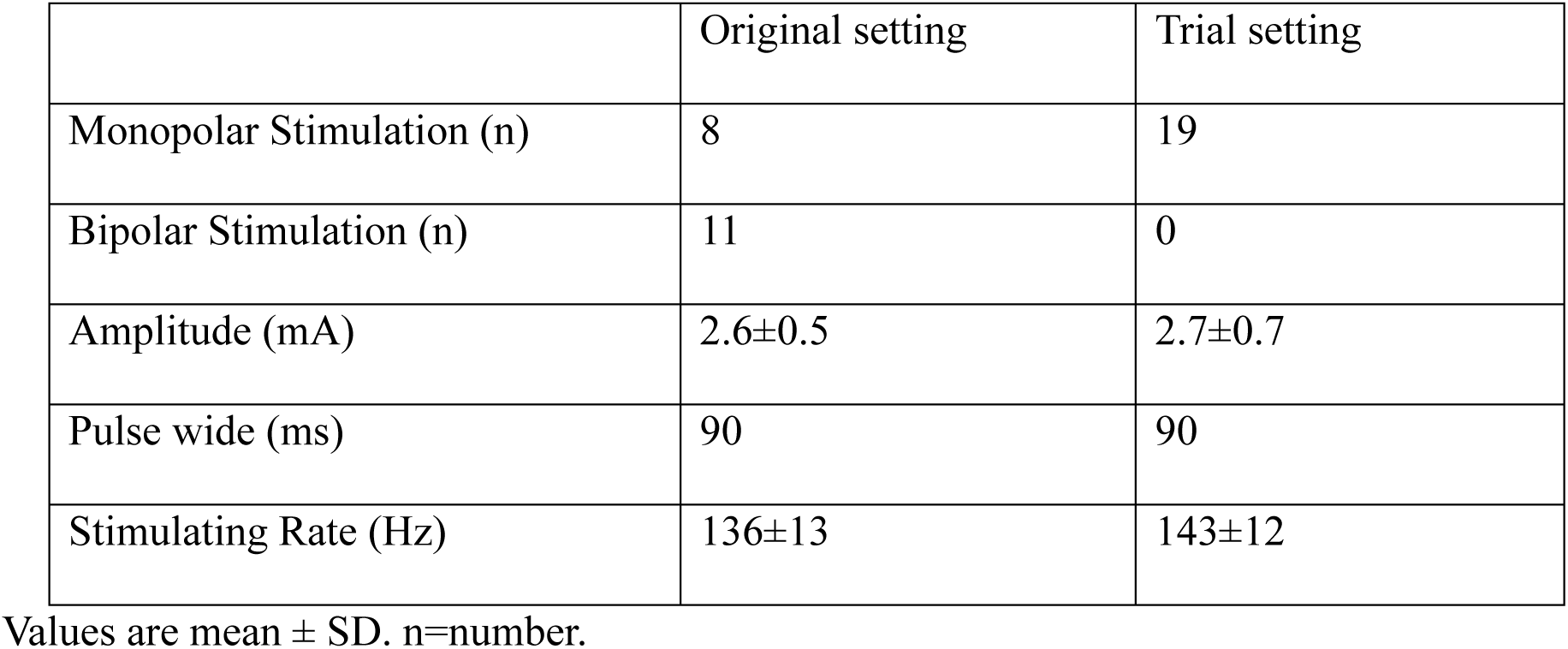
DBS parameters.

|  | Original setting | Trial setting |
| --- | --- | --- |
| Monopolar Stimulation (n) | 8 | 19 |
| Bipolar Stimulation (n) | 11 | 0 |
| Amplitude (mA) | 2.6±0.5 | 2.7±0.7 |
| Pulse wide (ms) | 90 | 90 |
| Stimulating Rate (Hz) | 136±13 | 143±12 |

Values are mean ± SD. n=number.

Three DBSs in three patients (two on the right side and one on the left) were identified as responders to the test stimuli, with a 10 mmHg increase in systolic blood pressure following stimulation. Random STM was then performed on the three responders for 12 minutes. The average STM-to-systolic AP transfer function exhibited low-pass characteristics with a corner frequency of 0.02 Hz. The average gain factor was 8.24 mmHg/STM at the steady state and decreased gradually as the input frequency increased. As shown in Figure 3, the phase spectrum revealed that the input-output relationship was in phase and that the phase delay increased towards higher frequencies (Figure 3).

**Figure 3.**
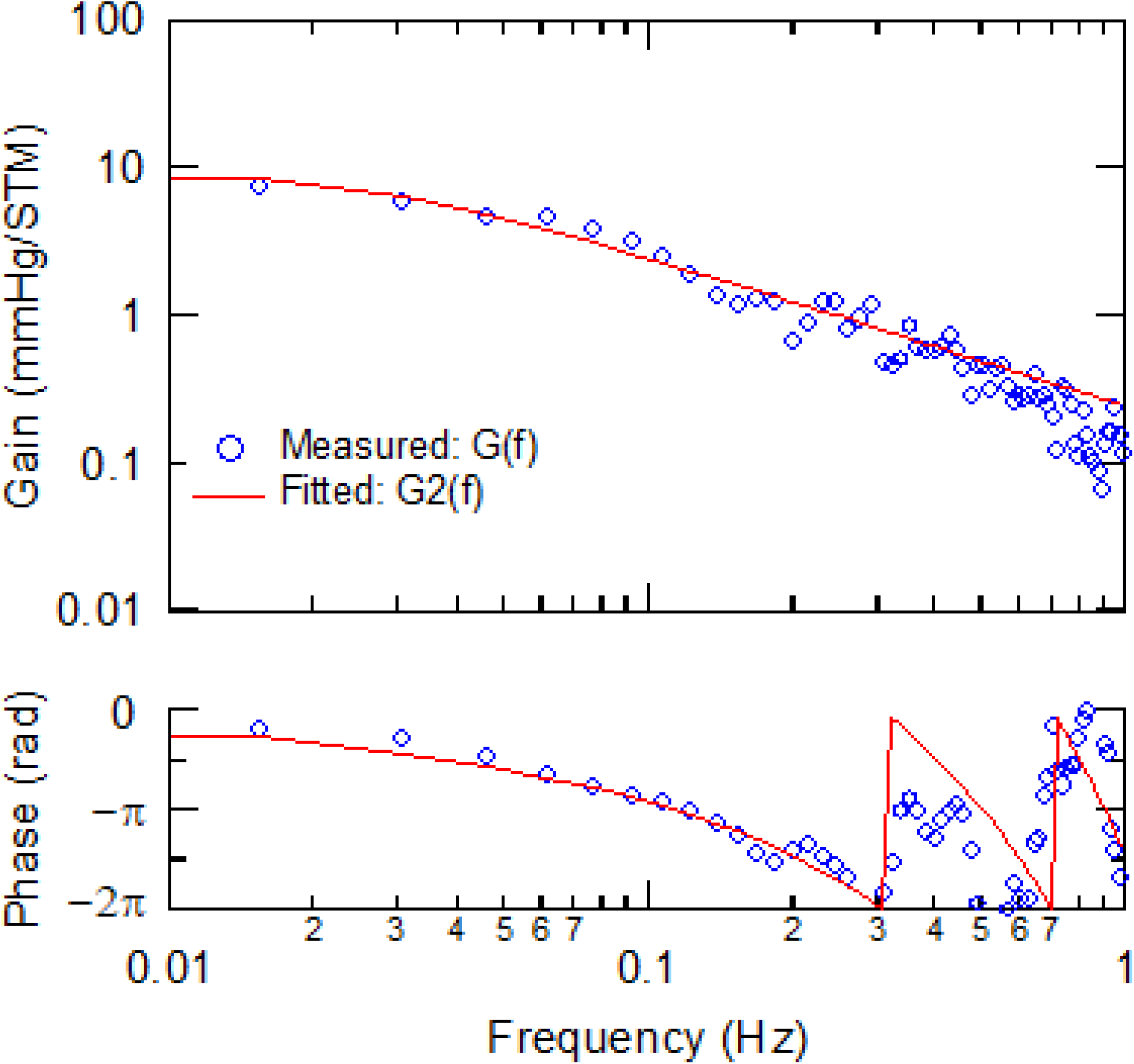
An average of the transfer function (G(*f*)) from DBS to AP in the 3 responders (upper panel) and the phase spectrum (lower panel). The averaged steady state gain was 8.24 mmHg/STM, and the gain decreased gradually with increasing input frequency.

The average of the estimated H₂(f) values from the three responders was used to simulate the design of the artificial vasomotor center. The adequate K_p_ was set at 0.12 and the K_i_ at 0.018. Computer simulations revealed that the system could quickly and effectively counteract a sudden 10 mmHg drop in blood pressure induced by an external disturbance, such as head-up tilting (Figure 4).

**Figure 4.**
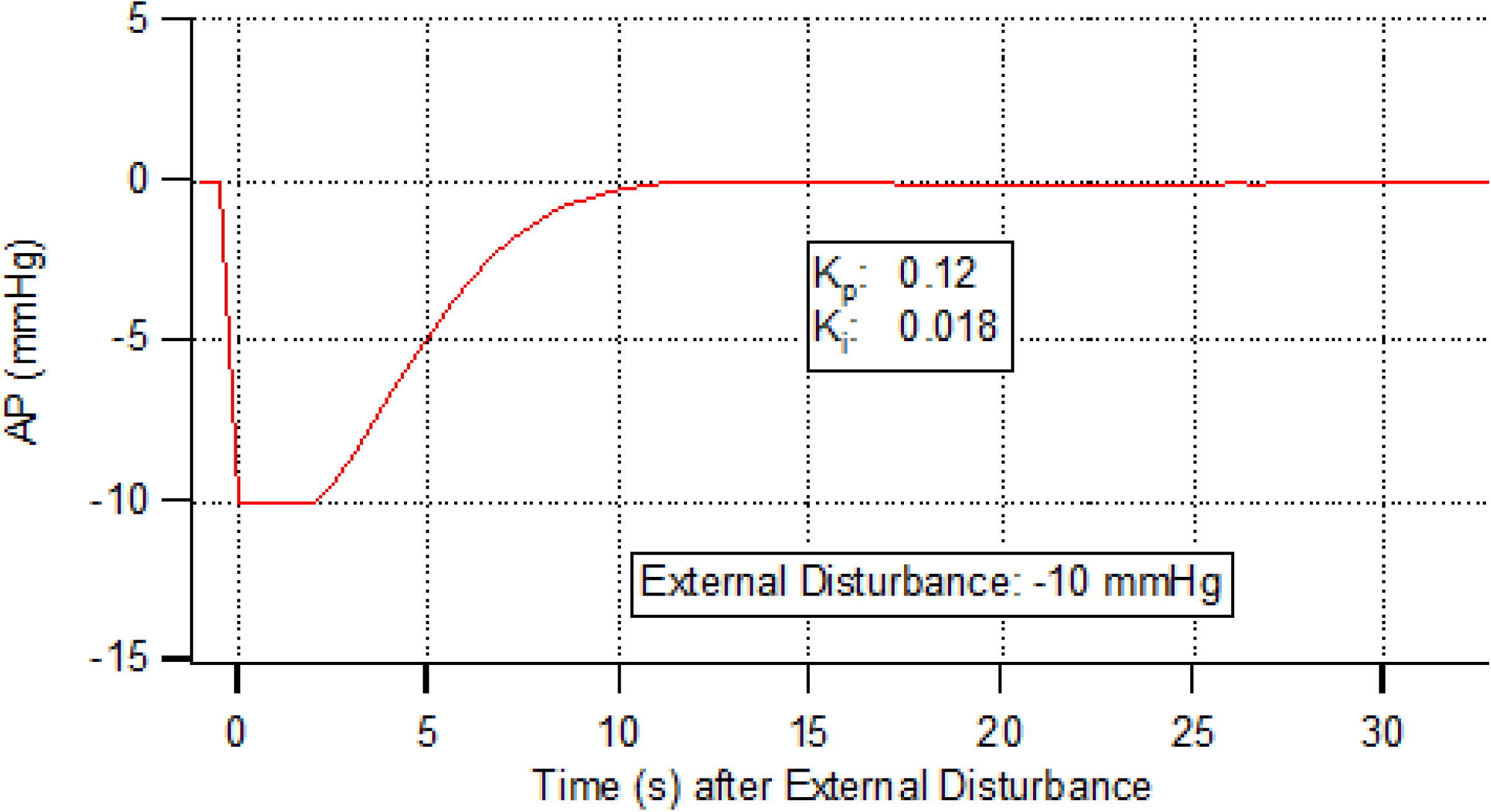
The results of simulations of a feedback system using the adequate parameters of K_p_:0.12 and K_i_:0.018 in H_2_(*f*). A stepwise pressure decline with an amplitude of 10 mm Hg is assumed to be imposed. The simulated artificial baroreflex with these parameters enabled the quick and stable minimization of the sudden AP drop induced by external disturbance.

## DISCUSSION

To evaluate the AP response to the STM of DBS, a white noise system identification approach was used in the present study. In contrast, earlier studies only observed and quantified AP responses to off-on or on-off changes in the STM (11–13). However, this traditional method has technical difficulties in accurately evaluating AP responses to STM. This is because the physiological system is rich in contaminating noise, which makes it difficult to identify the cause-effect relationship. For example, the AP should change or fluctuate spontaneously or in synchrony with cyclic vasomotion and breathing, independent of STM. Therefore, earlier studies may have underestimated or overestimated the effect of DBS on the AP, especially when spontaneous AP changes of unknown origin and the amplitude of Mayer waves and respiratory oscillations in the AP were much larger than the effect of DBS on the AP (19). The white noise system identification approach is largely unaffected by noise, providing a robust method for accurately estimating the input-output relationship.

The white noise method has another important advantage. Once the transfer function of the system is accurately identified in the frequency domain, the impulse–response function can be estimated. Thus, we can quantitatively visualize the transient response of the system using the impulse response function in the time domain and simulate the response to an external perturbation with any waveform under the closed-loop conditions of a negative feedback system, such as an artificial baroreflex system. Based on this theoretical background, the white noise system identification method was applied in this study.

The present study suggests the potential for creating an artificial baroreflex system by exploiting the effects of DBS on the AP. We have previously developed this method using electrical spinal stimulation (15,16). In this study, the gain factor of stimulation to mean arterial pressure was 0.43±0.13 mmHg/Hz at the steady state, and the feedback parameters in the servo system, with Kp set to 1 and Ki set to 0.1, could quickly and effectively attenuate the external disturbance to AP. In the present study, the steady-state gain from stimulation to systolic AP in the 3 responders was 8.24 mmHg/STM. The feedback DBS system attenuated the sudden AP drop of 10 mmHg induced by an external disturbance. This demonstrates the potential for developing a novel therapeutic system utilizing DBS. However, the gain in AP on stimulation in some responders was small. Therefore, we need an appropriate location for the electrodes in the STN and stimulation settings for effective modulation of the AP.

Several studies have shown the effects of STN-DBS on AP and autonomic systems (11–14), with STN-DBS on and off. Stimulation of the post-thalamic area of the periventricular and periaqueductal gray matter for chronic pain can affect the AP drop during head-up tilt standing (20,21). Stemper et al. showed that STN-DBS could maintain blood pressure while standing by turning bilateral STN stimulation on and off (11). They showed that the systolic AP dropped significantly from 133.9 to 116.2 mmHg when standing with DBS off, whereas there was no significant difference between 137.9 and 126.9 mmHg with DBS on. This indicates that STN-DBS prevented a drop in arterial pressure during standing. Conversely, some studies have reported that DBS is ineffective in the treatment of OH (22–25). The differences in these effects on the AP drop may be due to the settings of the DBS parameters or stimulation site. The stimulation site in the nuclei of the DBS was determined to be clinically effective for alleviating PD symptoms. The property settings and sites for OH may differ from those for PD symptoms.

The effects of the bilateral stimulation of the sympathetic nervous system and arterial pressure have been evaluated in several studies using STN-DBS (11–13). Cessation of bilateral stimulation can cause recurrence of symptoms and tremors. However, these symptoms and findings may increase the sympathetic activity. Therefore, we tested the effect of DBS on blood pressure using unilateral stimulation, excluding patients who experienced discomfort. In contrast, unilateral DBS of the periventricular and periaqueductal gray areas showed the same effects. Green et al. reported the beneficial effects of unilateral DBS on the AP and sympathetic activity (20,21). In the present study, we also showed that unilateral stimulation can affect the arterial pressure. The original electrode location and DBS settings were fixed to treat PD symptoms. Therefore, when bilateral stimulation is stopped, PD symptoms appear, which may stimulate the sympathetic nerves. We used unilateral stimulation to reduce symptoms as much as possible. Therefore, an additional stimulating position is required to sustain the reduction in arterial pressure and continue treating PD symptoms. A new system and generator are required to treat PD symptoms and AP drop simultaneously.

Although previous reports have indicated some anatomical differences in the effects of STN-DBS on the AP, the neural pathways involved in AP modulation in DBS are yet to be fully elucidated. Liu et al. demonstrated that the sympathetic index of low-frequency/high-frequency (LF/HF) heart rate variability was influenced by STN-DBS, with a negative correlation observed between LF/HF and the lateral electrode position from the mid-commissural point (26). They indicated that the pathway includes the STN and that the outflow tracts involve the zona incerta. Hyam et al. suggested that STN-DBS in patients with PD requires higher frequency stimulation than in other regions, and total energy delivery might stimulate proximity to the STN to indirectly stimulate the central autonomic network rather than the STN itself (27). In our study, no difference in the effect was observed at different stimulation frequencies or electrical intensities. Therefore, the intra-patient difference in the efficacy of STN-DBS for the AP might be due to the distribution of stimulating electrodes in the STN.

In conclusion, in the present study, we demonstrated that STN-DBS could restore arterial pressure during standing and during dynamic responses.

## Data Availability

The data that support the findings of this study are available from the corresponding author, FY, upon reasonable request.

## Acknowledgments

We thank Atsuko Murata and Yukari Yamamoto for their excellent technical assistance throughout this study.

## NOVELTY AND RELEVANCE

### What Is New?

Deep brain stimulation (DBS) often has a pressor effect through sympathoexcitation in the brain, indicating that the baroreflex failure in some patients with Parkinson disease (PD) is of a central type. This suggestion is associated with the applicability of bionic technology as a novel therapeutic strategy for orthostatic hypotension (OH) in PD. In the present study, we delineated the dynamic response of arterial pressure (AP) to DBS, devised a feedback-control algorithm for the vasomotor center of an artificial baroreflex system. We also demonstrated that subthalamic nucleus (STN)-DBS could restore arterial pressure during standing and the dynamic response.

### What Is Relevant?

In our previous study, we developed an artificial baroreflex system that electrically stimulated sympathetic nerves via an epidural catheter placed at the level of the lower thoracic spinal cord. This result prompted us to conclude that DBS could be an alternative to spinal cord stimulation and non-pharmacological therapy for OH in patients with PD.

### Clinical/Pathophysiological Implications?

A new artificial baroreflex system using DBS can restore arterial baroreflex function in patients with severe OH due to central baroreflex failure. This system can also be used in patients with OH and other neurological diseases apart from PD, including Shy-Drager syndrome and baroreceptor deafferentation.

## PERSPECTIVES

Using a white noise system identification approach, we designed an artificial vasomotor center to restore arterial baroreflex function. This system has the potential to serve as a new therapeutic modality for treating severe orthostatic hypotension (OH) in patients with Parkinson disease (PD) and various central baroreflex failure syndromes. Further studies with a larger number of patients with PD utilizing unified and variable stimulation settings and positions are required. Furthermore, the efficacy of this system for treating patients with OH should be evaluated in a clinical setting.

## Nonstandard Abbreviations and Acronyms

AP: arterial pressure
DBS: deep brain stimulation
ECG: electrocardiogram
OH: orthostatic hypotension
PD: Parkinson disease
STM: frequency of a pulse train
STN: subthalamic nuclei

